# Cystic Fibrosis Australia and Phage Australia survey: Understanding clinical needs and attitudes towards phage therapy in the CF community

**DOI:** 10.1101/2024.05.14.24307275

**Authors:** Stephanie Lynch, Holly Sinclair, Ameneh Khatami, Nicki Mileham, Jessica C Sacher, Jan Zheng, Ruby CY Lin, Jonathan Iredell

## Abstract

Cystic fibrosis (CF) is the most prevalent serious inherited disease in Australia, imposing significant health risks. CF is characterised by chronic lung inflammation and recurrent pulmonary infections that increase morbidity and premature mortality rates. The emergence of antimicrobial resistance (AMR) places further challenges on the treatment and management of CF, necessitating research into alternative strategies for treatment of bacterial infections. Bacteriophage therapy, involving bacterial-specific viruses, is a potential avenue for AMR infections in patients with CF. Existing literature supports the feasibility of phage therapy in CF but there has been a gap in investigating attitudes of the CF community including affected individuals and their caregivers, regarding phage therapy. Understanding perspectives and needs of the CF community is essential for successful implementation and acceptance of novel therapies including phage therapy.

We conducted a survey that encompasses responses from 112 consumers from across Australia, comprising people living with CF (38.4%), parents of affected children (49.6%), carers (6.4%), and family members (3%). The findings showed a significant reliance on antibiotics with 51.4% requiring oral, 43.4% nebulised, and 11.4% intravenous (IV) antibiotics within the preceding 2 weeks. Respondents highlighted the availability of new treatments, duration of hospitalisations and costs associated with treatment as important priorities to address. Despite an awareness of phage therapy among 62.4% of respondents, 86.4% expressed interest in obtaining more information, primarily from medical staff (66.7%). Notably, 96.0% of respondents expressed willingness to participate in phage therapy trials. The results of this survey highlighted the CF community’s strong interest in advanced therapeutic approaches, specifically phage therapy. The findings reveal a notable recognition and acceptance of phage therapy as a viable treatment option for pulmonary infections associated with CF.

## Introduction

Cystic fibrosis (CF) is one of the most prevalent inherited genetic disorders in Australia and it is estimated that the number of individuals living with CF will rise by 70.0% by 2025 in specific global regions ^1,2^. Data from the Australian CF Data Registry (ACFDR) currently records 3538 individuals living with CF, indicating an incidence of 1 in 2500 births ^3–5^. CF is a multi-organ condition primarily characterised by pulmonary infections leading to progressive reduction in lung function ^1,2^. In children living with CF, predominant pathogens are *Haemophilus influenzae* and *Staphylococcus aureus*, however, as lung disease progresses, patients become colonised with *Pseudomonas aeruginosa* and other Gram-negative non-fermenting bacteria ^6^. These pathogens frequently exhibit resistance to conventional antibiotics, necessitating prolonged courses of treatment that often do not achieve complete eradication ^7^. By adulthood, 80% of individuals become chronically colonised with *P. aeruginosa* ^*8*^. A 2016 Australian study found that 31.0% and 35.0% of sputum isolates were multi-antibiotic resistant *P. aeruginosa* (MARPA) among paediatric and adult patients with CF, respectively ^9^. Currently there is no cure for CF, and treatments, including antibiotics, aim to manage symptoms, prevent complications and improve quality of life. However, rising antimicrobial resistance (AMR) places this vulnerable group at even greater risk of morbidity and mortality from infections that are resistant to conventional antibiotics. This highlights the critical need for research into development of alternative methods to treat bacterial infections in patients with CF.

Phage therapy or the therapeutic administration of bacterio(phages), has re-emerged as an antimicrobial strategy^10^. Phages are bacterial viruses that recognise, attach to and replicate within selective bacterial targets ^11^. Due to this specificity to bacterial strains and inability to attack human cells, phages are generally regarded as safe. Phages can be used in cases where antibiotic options are limited or have associated toxicity ^12,13^. Numerous compassionate cases have administered phage to treat a diverse range of bacterial pathogens in patients following lung transplantation or in those with CF ^14–20^. These cases describe varying degrees of success in both microbiological and clinical outcomes, however, collectively support the safety and tolerability of phage therapy, without severe adverse events ^14–20^. Such reports highlight the potential of phage therapy as a viable approach to combat AMR bacterial infections in patients with CF. Despite these case reports ^14–20^ and the subsequent development of clinical trials for phage therapy in patients with CF ^21,22^, clinical priorities of the CF community (patients and carers) and their attitudes toward phage therapy has yet to be explored.

To note, Australians have access to universal healthcare and patients with CF have access to subsidised and specialised disease management plans (https://www.cysticfibrosis.org.au/). This study aimed to survey both people living with CF and the broader CF community in Australia, including carers and family members of people with CF to understand the current burden of antimicrobial therapy in this population, and their perspectives regarding clinical trials involving phage therapy. Ultimately, the survey results could be used to develop an advocacy framework for phage therapy to be presented to the Therapeutic Goods Administration of Australia. It also seeks to realign the priorities of phage therapy centres like Phage Australia by utilising a co-designed framework to advance phage therapy specifically for CF.

## Methods

### Survey design and distribution

An anonymous online research survey was developed to understand the clinical needs of individuals and carers within the CF community in Australia, in particular, their attitudes toward clinical trials using new treatments, such as phage therapy. As the survey was anonymous and open to the public, completion of the survey was considered implied consent to participate in the survey. The survey questions were designed in a close-ended format to facilitate statistical analysis using descriptive statistics. A team of clinicians, with expertise in caring for CF patients and experience in successfully treating patients with phage therapy, formulated the survey questions. In addition, these clinicians actively engage in phage therapy research. We also engaged Cystic Fibrosis Australia (CFA) in survey design, specifically framing questions to increase engagement. Thirteen single choice questions were input into Survey Monkey (https://www.surveymonkey.com). The survey could be completed in approximately 2 minutes.

The CF community was provided with a digital survey link through the Cystic Fibrosis Australia ‘Fight Fire with Fire’ newsletter (Supplementary 1). The newsletter also contained a brief introduction to phage therapy and to Phage Australia, to give context prior to completing the survey. The survey remained open for 4 months, aiming for at least 100 respondents (Supplementary 2).

The data was automatically collated on Survey Monkey, a password-protected platform accessible exclusively to study investigators. No identifiable information was collected. Participants had the opportunity to review responses prior to submission and modify before submission to ensure their comfort with participation and accuracy of data collected.

### Data analysis and visualisation

Survey responses collated from Survey Monkey were downloaded into a password-protected CSV file. GraphPad Prism software was utilised where responses were visualised, including bar graphs depicting response percentages.

## Results

### Demographics of survey participants

**Table 1** represents the demographic characteristics of the 112 participants. Geographically, there was a notable concentration of responses from individuals in different state in Australia namely, NSW (New South Wales), VIC (Victoria), and WA (Western Australia), indicating a higher participation rate from these regions.. None of the respondents (98.4%) identified themselves as having Aboriginal or Torres Strait Islander ancestry. Of the 112 participants, 71.2% identified as female. Age distribution showed the majority (44.0%) falling within the 40-59 age group, followed by the 20-39 age groups (31.2%). This indicates a diverse representation across different age brackets within the surveyed population.

**Table 1.** Demographics of the 112 respondents of the survey.

|  | Number of responses (% of responses). Total responses n =112 |
| --- | --- |
| **Respondent CF status** |  |
| Respondent has been diagnosed with CF | 48 (36.40%) |
| Respondent is a parent of a child with CF | 62 (49.60%) |
| Respondent is a family member of someone with CF (e.g. sibling, aunt/uncle) | 8 (6.40%) |
| Other | 4 (3.20%) |
| **Respondent age demographics** |  |
| 13-19 | 20 (16.00%) |
| 20-39 | 39 (31.20%) |
| 40-59 | 55 (44.00%) |
| 60-99 | 14 (11.20%) |
| **Aboriginal and Torres Strait Islander origin** |  |
| None of the above | 123 (98.40%) |
| Yes, Aboriginal | 1 (0.80%) |
| Yes, Torres Strait Islander | 1 (0.80%) |
| Yes, both Aboriginal and Torres Strait Islander | 0 (0.00%) |
| **Gender demographics** |  |
| Male | 35 (28.00%) |
| Female | 89 (71.20%) |
| Non-binary | 1 (0.80%) |
| **Geographical location** |  |
| Victoria (VIC) | 25 (20.00%) |
| New South Wales (NSW) | 31 (24.80%) |
| Australian Capital Territory (ACT) | 11 (8.80%) |
| Queensland (QLD) | 13 (10.40%) |
| Northern Territory (NT) | 1 (0.80%) |
| Western Australia (WA) | 24 (19.20%) |
| South Australia (SA) | 12 (9.60%) |
| Tasmania (TAS) | 6 (4.80%) |

### Antibiotic reliance and clinical priorities among CF patients

As shown in **Figure 1**, within the preceding two weeks, 51.4% required oral, 43.3% nebulised, and 11.4% intravenous (IV) antibiotics. Some respondents indicated concurrent use of oral, nebulised, or IV antibiotics within this two-week period.

**Figure 1.**
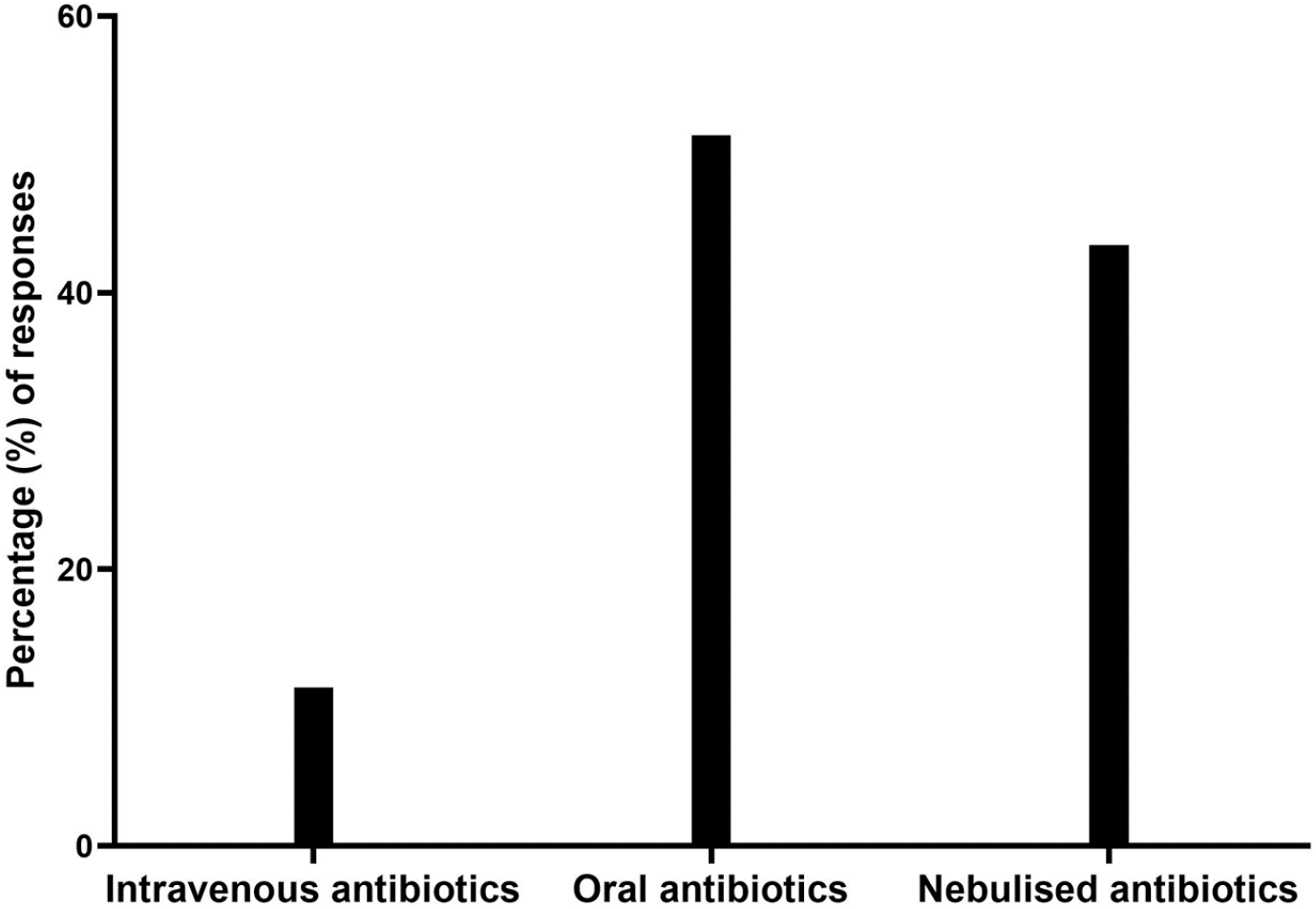
Recent (past 2 weeks) use of antibiotic modalities for the treatment or management of CF.

Almost half (43.9%) of respondents reported moderate antibiotic use (1-3 courses per year) to manage CF symptoms, while a quarter indicated infrequent use (<1 course per year) and a similar proportion indicating 3-6 courses per year were used (Figure 2). Response also revealed that the majority (67.9%) of participants required less than one course of intravenous antibiotics to be given in hospital, annually.

**Figure 2.**
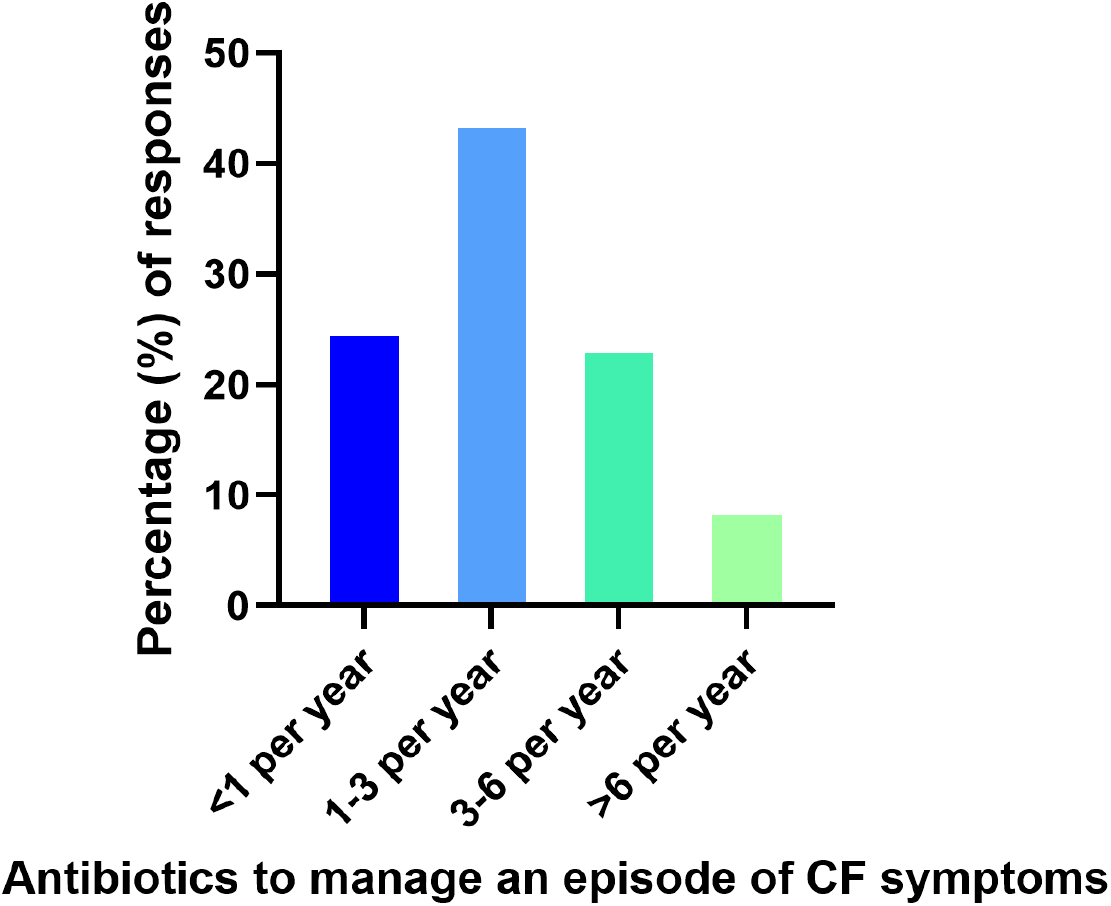
Annual frequency of antibiotic courses for managing acute episodes of CF. Responses exclude the long-term prophylactic antibiotics taken.

The greatest priority among 80.80% of survey participants was placed on availability of new treatments (**Figure 3**). Subsequently, time spent in hospital was a concern for 12.0% of participants. A smaller proportion of participants expressed concerns about the cost of therapies (6.4%) and communication from researcher to patients (0.80%).

**Figure 3.**
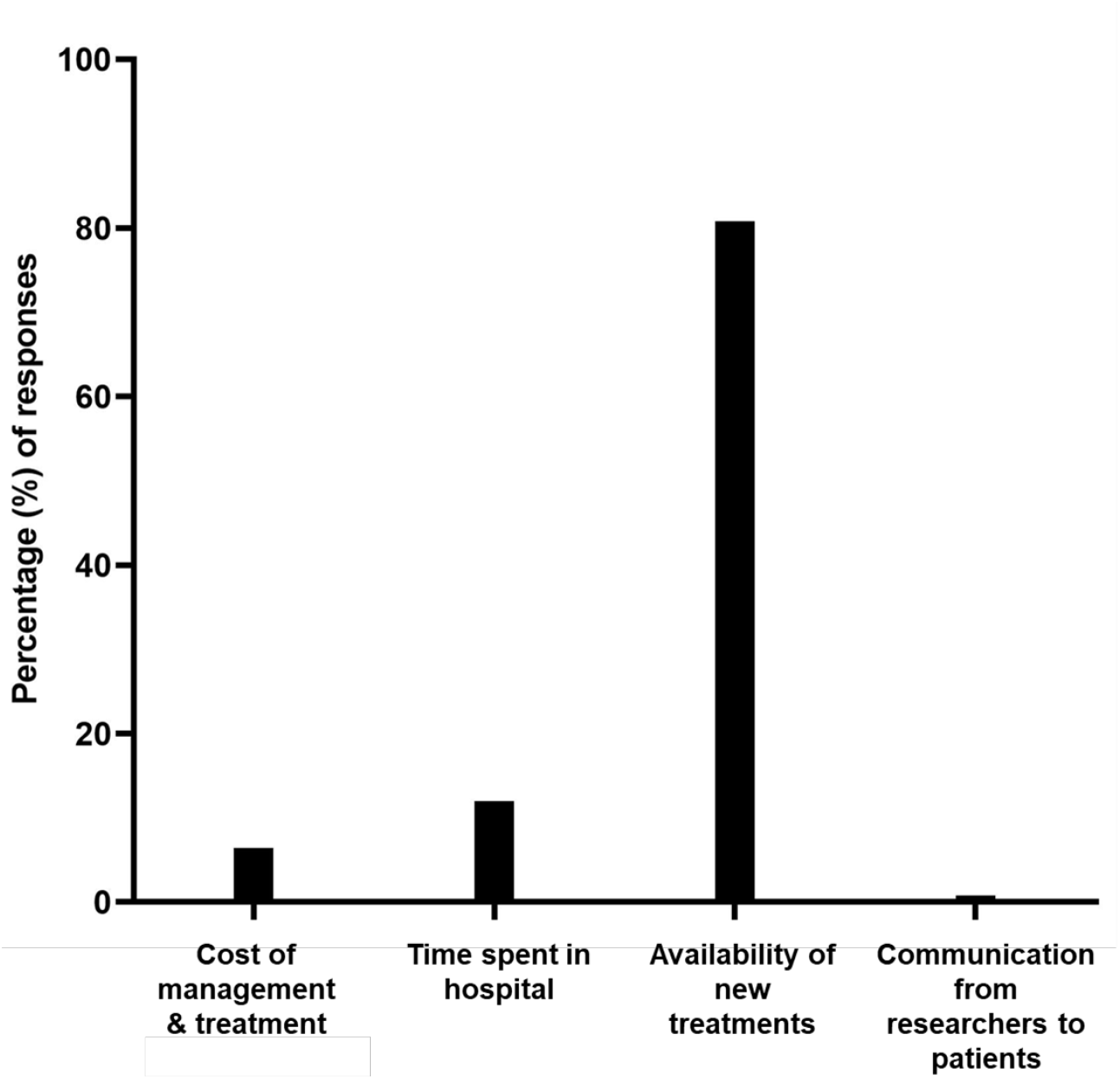
Priorities to be addressed for the management and/or treatment of CF based on responses from the CF community.

### Awareness, engagement, and uptake of phage therapy within the CF community

The majority of respondents (62.4%) indicated an awareness of phage therapy, recognising its potential as an alternative or adjunctive treatment to antibiotics for infections in CF.

Furthermore, majority of survey participants (86.4%) indicated interest in learning more about phage therapy as an adjunct to antibiotics for CF treatment and/or management. Among those, the majority (66.7%) indicated they would rely on medical staff for more information (**Figure 4**).

**Figure 4.**
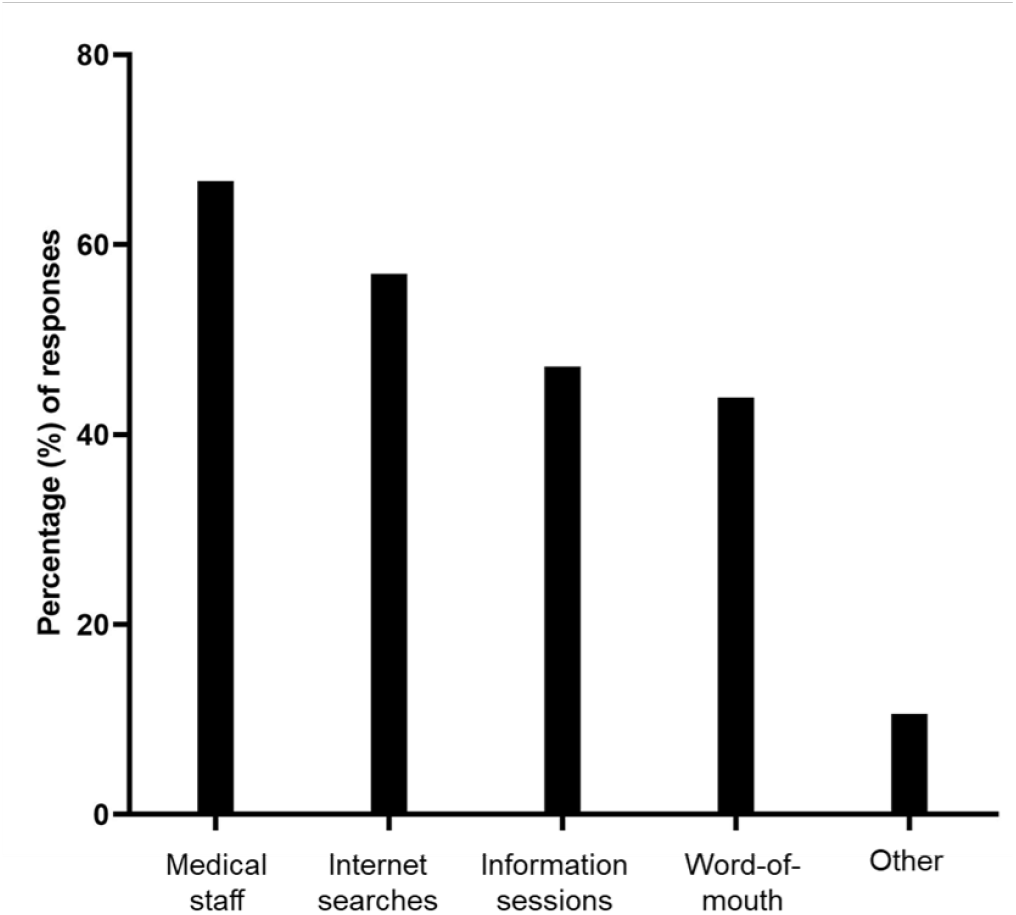
Potential avenues the CF community would use to find out more about phage therapy.

An overwhelming majority (96.0%) of respondents expressed willingness to participate in a clinical trial investigating phage therapy. Similarly, 95.2% indicated an increased inclination towards engaging in phage therapy trials if government subsidised.

## Discussion & Conclusions

In response to limited knowledge about the CF community’s perception of emerging therapies, such as phage therapy, we conducted a survey to bridge this gap. Despite the longstanding use of phage therapy in medicine, and in treating infections in patients with CF, there has been limited exploration of this area. Our survey aimed to provide valuable insights into the CF community’s perspectives on this issue.

The demographics of the respondents of this survey align with recent summaries of the Australian CF Data Registry (ACFDR), highlighting a median age of 20.2 years among individuals with CF, with children (<18 years) constituting 44.5% of the CF population ^5^, consistent with the high proportion (almost 50%) of parent survey respondents. This highlights high engagement of parents among the CF community, and the importance of clinical trials to include paediatric populations. Geographic representation of survey responses was also largely consistent with ACFDR data on States of residence for individuals with CF (NSW = 1014 people, QLD = 908 and VIC = 724), although there was a lower-than-expected response rate from Queensland ^5^.

Various pathogens, including *P. aeruginosa, H. influenzae, S. aureus*, and *Mycobacterium*, are associated with chronic infections in CF, leading to high antibiotic use in this population, either for prophylaxis or to manage exacerbations ^23,24^, and subsequent risk of developing multi-drug-resistant infections. Our survey responses confirmed this, with over half reporting oral antibiotic use within the preceding two weeks. Almost half of the participants reported requiring 1-3 courses of antibiotics per year, although the majority required less than one course (of presumably intravenous antibiotics) in hospital. These data are consistent with a previous study showing the majority of pulmonary exacerbations in patients with CF are managed with oral rather than intravenous antibiotics, particularly in young children ^29^. Interestingly, a study investigating the impact of TRIKAFTA® (Vertex Pharmaceuticals, Boston, United States of America; elexacaftor/tezacaftor/ivacaftor) initiation on infection-related medical visits in patients with CF showed a significant and rapid decrease in antibiotic use following TRIKAFTA^®^ initiation ^25^. A limitation of our survey is that Trikafta use was not specifically enquired about, which could have provided valuable stratification of the data. Nebulised antibiotics were utilised by just under half of respondents consistent with other data indicating an increased use of long-term inhaled antibiotics in patients with CF ^25^. Overall, the high burden of antibiotic use to manage exacerbations of CF symptoms was evident from survey responses, highlighting the significant impact chronic and recurrent chest infections have on the quality of life of patients with CF. Consequently, the survey results indicated a significant majority of respondents prioritised accessibility of new treatments for CF indicating an urgent need for alternative approaches to management of infections in patients with CF, particularly amidst the growing AMR crisis.

One advanced therapeutic approach already in use is phage therapy, the targeted use of phages against bacterial pathogens. Interestingly, over half of respondents had prior knowledge of phage therapy, in contrast to a 2023 study surveying the UK general public where only 13% were aware of it ^26-28^. This suggests that people with CF or their families may be more informed about novel therapies. Additionally, our survey found there was a strong interest among respondents in learning more about phage therapy, with the majority selecting medical staff as their primary information source. However, it is important to note that medical professionals themselves may require comprehensive education on phage therapy including data from clinical trials, as 42.0% of Australian infectious disease physicians and 38.5% of Korean infectious disease physicians surveyed reported only moderate knowledge about phage therapy ^29,30^. A high level of willingness to participate in phage therapy trials was also expressed by survey respondents, with an even higher level of willingness to pursue new therapies if the government provided subsidies for treatment costs. This may reflect existing economic burdens associated with healthcare placed on this vulnerable population. To date, experience in using phage therapy for CF infections primarily comprises compassionate access cases ^14–20^. Thus, there is a critical need for increased access to clinical trials to systematically examine the effectiveness of phage therapy, however, only six clinical trials involving phage therapy and patients with CF are currently registered on clinicaltrials.gov ^22^.

The survey results provide valuable consumer insights for regulatory bodies in regard to antibiotic usage, and preferences for phage formulation and routes of administration. Data from clinical trials are needed to determine the most effective approaches, including the role of adjunctive antibiotics, duration of therapy and the efficacy of single phage therapy versus a cocktail approach ^31^.

In addition, the survey results reflected that the key priorities of CF patients remain the same, i.e., management of respiratory health including adherence to prescribed medication, reduced reliance on antibiotics, and managing airway clearance and avoidance of respiratory infections. Utilising phage therapy to improve quality of life that include shortened hospital stays and ameliorated daily functioning is implied.

This survey highlighted the importance of establishing a formalised knowledge-sharing network to educate and inform consumers, healthcare providers, scientists and regulatory bodies about phage therapy. These measures can potentially accelerate the availability of phage therapy to individuals with cystic fibrosis.

## Supporting information

Supplementary 1 Survey flyer

Supplementary 2 Survey questions

## Data Availability

All data produced in the present study are available upon reasonable request to the corresponding author

## Acknowledgments

We would like to thank Nettie Burke, former CEO of Cystic Fibrosis Australia for her support and advice throughout this study. This work was funded by Australia’s Medical Research Future Fund, Frontiers Stage 1 (RFRHPI000017) to Iredell, Lin, Khatami *et al*., 2021: Phage Australia - Integrating Australian Phage Biobanking and Therapeutic Networks and Delivering Solutions for Antimicrobial Resistance.

## Authors’ contribution

Conception: JI, RL

Coordination: RL, SL, NM

Question design: JI, RL, AK, HS, SL, NM, JS, JZ

Data analysis: SL

Manuscript: SL, HS, RL, AK

