## Supplementary 1 Survey flyer for "Cystic Fibrosis Australia and Phage Australia survey: Understanding clinical needs and attitudes towards phage therapy in the CF community"

Subscribe | Past Issues | Translate

View this email in your browser

9 November 2021

### FIGHTING FIRE WITH FIRE

In both the CF Community and society at large, we are used to thinking of bacteria as the enemy. A great deal of medical care revolves around curtailing the influence of invisible microscopic interlopers. But the future of medicine may just lie with a naturally occurring virus that kills only bacteria known as bacteriophages, ‘phages’ for short.

At a time when society has invested heavily into antibiotics and vaccines (for good reason), bacteriophages are a promising avenue for fighting disease and the rise of antimicrobial resistance.

People with CF will be among the primary beneficiaries, because CF heroes like Jon Iredell and Ruby Lin are working hard behind the scenes to adapt bacteriophage technology to fight certain infections common to our community.

Phage therapy is indeed already in use in certain forms of CF treatment, but each phage treatment needs to be precisely and painstakingly targeted to a specific bacteria that it is being used to fight. This means that more research and better data is called for if we are going to get the most out of this technology.

We need amazing researchers like Jon Iredell and Ruby Lin working to develop these phages, but right now their team (Phage Australia) needs us. Help her latest project by clicking HERE and completing the survey. We want to get the word out about Phage therapy and make it part of the national conversation around health. This technology has too much promise and potential to be ignored or sidelined.

Bacteriophages turn the threat back against our most deadly microbial enemies,

The Team at CFA

***CFA Wine Drive***
http://www.labelmywine.com.au/winedrives/cystic-fibrosis-65-roses/

***Entertainment Books and Entertainment Digital Memberships***
https://www.entertainmentbook.com.au/orderbooks/9444e64

*Copyright © 2021 Cystic Fibrosis Australia, All rights reserved.*

Want to change how you receive these emails?
You can update your preferences or unsubscribe from this list.
