## Supplementary 2 Survey questions for "Cystic Fibrosis Australia and Phage Australia survey: Understanding clinical needs and attitudes towards phage therapy in the CF community"

Copy of PHAGE THERAPY IN THE CF COMMUNITY

**Target audience to receive the survey:**
**Cystic fibrosis (CF) patients directly or general CF community (e.g. carers, siblings, other family members).**

**Target audience to receive the survey reports/outcomes:**
**TGA, CF community & Phage Australia.**

**Key messages/outcomes from the survey:**
- **TGA: Propose an advocacy strategy for phage therapy in the CF community and assess payment pathway.**
- **CF Community: New therapy availability.**
- **Phage Australia: How to proceed with advocacy to TGA and inform how to design payment paths.**

**What is Phage Therapy?**
**Modern medicine has adopted two distinct and complementary approaches to infectious disease control. Vaccination can prevent infection – but is only available for a small number of infectious agents. For infections that are not prevented, antimicrobials (mostly antibiotics, with a few antivirals and antifungals) offer the possibility of treatment.**

**Antibiotics are failing due to growing antimicrobial resistance (AMR) and the R&D pipeline for new antibiotics has all but dried up. As the World Health Organization has warned, this threatens "an end to modern medicine as we know it". Common infections will once again become untreatable.**

**Phage therapy offers a third approach to infectious disease control. Phages (also known as bacteriophages) are viruses that prey on bacteria. They remain completely effective against antibiotic resistant bacterial strains, offering a last defence against otherwise untreatable infection.**

**Phage therapy also brings a number of critical advantages over antibiotic treatment. With careful preparation to remove impurities, they are non-toxic to humans. Unlike antibiotics, each phage is highly precise in the specific bacteria that it targets, meaning that treatment has fewer effects on the healthy bacteria in our bodies. They can be used on their own or in combination with other phages and antibiotics to increase efficacy even more.**

**Phage therapy has been successfully implemented in CF cases under the Special Access Scheme through Therapeutic Goods Administration at the Children's Hospital, Westmead and Westmead Hospital for *Mycobacterium abscessus* infection.**

**Thank you for taking part in the survey to help Phage Australia better understand the clinical impacts and needs of CF patients. Responses of this survey will help inform actions and payment pathways of upcoming therapies. This survey should take approximately 10 minutes.**

* **Q1**. Have you or someone you know been diagnosed with cystic fibrosis (CF)?

Other (please specify)

* **Q2.** Demographics - for CF patients. Aboriginal or Torres Strait Islander patients (and carer answering on behalf of the patient).

a) Your age group:

- ☐ 13-19
- ☐ 20-39
- ☐ 40-59
- ☐ 60-99

* b) Are you of Aboriginal or Torres Strait Islander origin?

- ○ None of the above
- ○ Yes, Aboriginal
- ○ Yes, Torres Strait Islander
- ○ Yes, both Aboriginal and Torres Strait Islander

* c) Gender

- ○ Male
- ○ Female
- ○ Non-binary
- ○ Self specified:

* d). Where are you located?

- ○ VIC
- ○ NSW
- ○ ACT
- ○ QLD
- ○ NT
- ○ WA
- ○ SA
- ○ TAS
- ○ Other (please specify)

* **Q3.** In the last two weeks have you/someone you care for used any of the following for the treatment or management of CF?

| | Yes | No |
|---|---|---|
| Intravenous antibiotics | ○ | ○ |
| Oral antibiotics | ○ | ○ |
| Nebulised antibiotics | ○ | ○ |

* **Q4.** To measure treatment impact, please indicate the following in respect to this year (2021):

| | <1 per year | 1-3 per year | 3-6 per year | >6 per year |
|---|---|---|---|---|
| Courses of antibiotics to manage an episode of worsening of CF symptoms (Please do not include long-term prophylactic antibiotics that you take regularly) | ☐ | ☐ | ☐ | ☐ |
| IV antibiotics given in hospital | ☐ | ☐ | ☐ | ☐ |
| IV antibiotics at home only (not following a short stay in hospital) | ☐ | ☐ | ☐ | ☐ |

* **Q5.** If one thing could be changed in the treatment/management of CF, what would be your priority? (Tick one that would be your top priority).

- ☐ Cost of management and treatment therapies
- ☐ Time spent in hospital
- ☐ Availability of new treatments
- ☐ Communication from researchers to patients

* **Q6**. Have you heard about Phage Therapy as an alternative or combination treatment to antibiotics for CF?

- ◯ Yes
- ◯ No

* **Q7.** Do you want to know more about Phage Therapy as an adjunct to antibiotics for CF?

- ◯ No - move to Q8
- ◯ Yes - please answer below

*As you were interested finding out more information about Phage Therapy, how would you generally find out more information about new treatments?*

- ☐ Medical staff (specialists, doctors, nurses)
- ☐ Internet searches
- ☐ Information sessions (organised by CFA, hospitals or universities)
- ☐ Word-of-mouth within CF community
- ☐ Other (please specify)

* **Q8.** If you felt properly informed and your usual doctor recommended it, would you or the person you are caring for consent to trial phage therapy as a treatment or alternative or combination to antibiotics?'

- ◯ Yes
- ◯ No

* **Q9.** Would you or the person you are caring for be more willing to trial phage therapy if the government were to subside the cost of treatment?

- ◯ Yes
- ◯ No

**Thank you for taking part in the survey. For more information surrounding the use of phage therapy head to https://criticalinfection.com/phage-australia/**

**CFA in collaboration with Phage Australia would like to publish the results of this survey. Any personal details will remain private and secure. Deidentified data will be analysed and you will be informed of the outcome. If you do not want your response of this survey to be included in this data analyses please.**
